# Replication and cross-validation of T2D subtypes based on clinical variables: an IMI-RHAPSODY study

**DOI:** 10.1101/2020.12.21.20248628

**Authors:** Roderick C Slieker, Louise A Donnelly, Hugo Fitipaldi, Gerard A Bouland, Giuseppe N. Giordano, Mikael Åkerlund, Mathias J. Gerl, Emma Ahlqvist, Ashfaq Ali, Iulian Dragan, Andreas Festa, Michael K. Hansen, Dina Mansour Aly, Min Kim, Dmitry Kuznetsov, Florence Mehl, Christian Klose, Kai Simons, Imre Pavo, Timothy J. Pullen, Tommi Suvitaival, Asger Wretlind, Peter Rossing, Valeriya Lyssenko, Cristina Legido Quigley, Leif Groop, Bernard Thorens, Paul W Franks, Mark Ibberson, Guy A Rutter, Joline WJ Beulens, Leen M ’t Hart, Ewan R Pearson

## Abstract

**Aims/hypothesis:** Five clusters based on clinical characteristics have been suggested as diabetes subtypes: one autoimmune and four subtypes of type 2 diabetes (T2D). In the current study we replicate and cross-validate these T2D clusters in three large cohorts using readily measured variables in the clinic.

**Methods:** In this cross-sectional study, 15,940 individuals were clustered based on age, BMI, HbA1c, random or fasting C-peptide and HDL in three independent cohorts. Clusters were cross-validated against the original clusters based on HOMA measures. In addition, between cohorts, clusters were cross-validated by re-assigning people based on each cohort’s cluster centres.

**Results:** Five distinct T2D clusters were identified and mapped back to the original four ANDIS clusters. Using C-peptide and HDL instead of HOMA-B and HOMA-S three of the clusters mapped with high sensitivity (80.6 – 90.7%) to the previously identified Severe Insulin Deficient (SIDD), Severe insulin resistant (SIRD) and Obese (MOD) clusters. The previously described ANDIS MARD cluster could be mapped to the two milder groups in our study – one characterised by high HDL, and the other having not any extreme characteristic (MDH cluster). When these two milder groups were combined they mapped well to the previously labelled MARD cluster (sensitivity 79.4%). In the cross-validation between cohorts, particularly the SIDD and MDH cluster cross-validated well with sensitivities ranging from 73.3% to 97.1%. SIRD and MD showed a lower sensitivity ranging from 36.1% to 92.3% where individuals shifted from SIRD to MD and vice versa.

**Conclusions/interpretation:** Clusters based on C-peptide instead of HOMA measures result in clusters that resemble those based on HOMA measures, especially for SIDD, SIRD and MOD. By adding HDL, the MARD cluster based upon HOMA measures resulted in the current clustering in two clusters with one cluster having high HDL levels. Cross-validation between cohorts showed generally a good resemblance between cohorts. Together, our results show that the clustering based on clinical variables readily measured in the clinic (age, HbA1c, HDL, BMI and C-peptide) results in informative clusters that are representative of the original ANDIS clusters and stable across cohorts.

## INTRODUCTION

A recent study stratified people with any form of diabetes into five clusters based on six clinical variables, i.e. age, glutamate decarboxylase (GAD) antibodies, BMI, HbA1c, insulin resistance (HOMA2-IR) and β-cell function estimates (HOMA2-B).[1] The five clusters were characterized by autoimmunity (SAID), insulin deficiency (SIDD), insulin resistance (SIRD), high BMI (MOD) and the last without any extreme characteristics other than high age (MARD).[1] Clustering of people with diabetes has been repeated successfully in several other studies based on these variables in people from European descent, other ethnicities and based on different clinical measures.[2-9] In addition, the original and subsequent papers have shown that people in different clusters have different risks for a number of diabetes related outcomes.[1-4] The autoimmunity and insulin deficient clusters were defined by high HbA1c at diagnosis and had higher risk on ketoacidosis and retinopathy[2, 7] and progressed more rapidly onto insulin relative to the other clusters.[1] Moreover, a recent study comprising multiple cohorts enriched for cardiovascular risk assigned people to the clusters identified by Ahlqvist et al.[1] based on the distance to the respective cluster centres. In this study, people in the SIDD cluster showed higher risk of MACE.[5] For the insulin resistant cluster a higher frequency of non-alcoholic fatty liver disease has been observed and people in this group were at increased risk of developing chronic kidney disease.[1] As HOMA calculations require fasting insulin or c-peptide and fasting glucose their measurement is not routine in clinical practice.

The aim of the current study is to perform a systematic replication and cross-validation of clustering based on five routine clinical variables in three large international cohorts (DCS, ANDIS, GoDARTS). In ANDIS, we directly compare the current clustering with those identified in the original study.[1]

## METHODS

### Cohort descriptions

Data from 15,940 individuals from three cohorts, DCS (Netherlands), GoDARTS (Scotland) and ANDIS (Sweden) were used in this cross-sectional study. Inclusion criteria for RHAPSODY were age of diagnosis was ≥35, clinical data available within 2 years after diagnosis, GAD negative, no missing data in one of the five for clustering used clinical measures and the presence of GWAS data.

#### Hoorn DCS cohort

The Hoorn Diabetes Care System (DCS) cohort is an open prospective cohort started in 1998 with currently over 14,000 individuals with T2D from the northwest part of the Netherlands.[10] The study has been approved by the Ethical Review Committee of the VU University Medical Center, Amsterdam. People visit DCS annually to monitor their diabetes. During this visit multiple measurements are collected as part of routine care, including anthropometric- and lab measurements. Measurements were used anonymously. Individuals were informed about the use of their data and were offered an opt-out. All laboratory measurements were done on samples taken in a fasted state. HbA1c measurements were performed using the turbidimetric inhibition immunoassay for haemolyzed whole EDTA blood (Cobas c501, Roche Diagnostics, Mannheim, Germany, run CV 1.6%).[10] HDL (mmol/L) was measured enzymatically (Cobas c501, Roche Diagnostics). C-peptide was measured on a DiaSorin Liaison (DiaSorin, Saluggia, Italy). In total, 2,953 individuals matched the inclusion criteria.

#### GoDARTS

For clinical purposes, individuals with diabetes mellitus from the Tayside region of Scotland (N□=□391,274; January 1996) were added to the DARTS register.[11] Retrospective and prospective longitudinal anonymized data were collected, including data on prescribing, biochemistry, and clinical data. All laboratory measurements were measured in a non-fasted state. People with T2D were asked to participate in the Genetics of Diabetes Audit and Research Tayside Study (GoDARTS), which currently includes over 10,000 individuals with T2D.[11] The GoDARTS study was approved by the Tayside Medical Ethics Committee. Informed consent was obtained from all participants. C-peptide was measured on a DiaSorin Liaison (DiaSorin, Saluggia, Italy). In total, 5,509 individuals matched the inclusion criteria.

#### ANDIS

The All New Diabetics in Scania (ANDIS) cohort aims to recruit all people with incident diabetes within Scania County, Sweden. Recruitment started in January 2008 until November 2016. People are included in the study close to diagnosis, with a median of 40 days (IQR 12-99). All laboratory measurements were measured in a fasted state. HbA1c measurements were obtained from the Clinical Chemistry database. C-peptide were determined with an electro-chemiluminescence immunoassay on Cobas e411 (Roche Diagnostics, Mannheim, Germany) or a radioimmunoassay (Human C-peptide RIA; Linco, St Charles, MO, USA; or Peninsula Laboratories, Belmont, CA, USA). In total, 7,478 individuals matched the inclusion criteria.

### Statistical analysis

Clustering was performed on five risk factors for type 2 diabetes progression [12], age (years), BMI (kg/m^2^), HbA1c (mmol/mol), HDL (mmol/L) and C-peptide (nmol/L). The latter two were included as proxies of beta-cell function and insulin sensitivity in absence of fasting glucose (GoDARTS) and insulin (DCS and GoDARTS) and therefore HOMA measures in these cohorts in GoDARTS. K-means clustering was performed separately for males and females using the *kmeansruns* function in the R-package *fpc*. The optimal number of clusters was determined using the gap statistic across the three cohorts,[13] this being defined as the point where the curve of the GAP statistic versus the number of clusters flattened, with little added value of increasing the number of clusters. The stability of the clusters was assessed in two ways. The clusters identified here in ANDIS using C-peptide instead of HOMA were compared to their previously published clusters based on HOMA.[1] Second, identified clusters were cross-validated between cohorts to assess their stability. For this, individuals from cohort A were assigned to clusters based on the cluster centres of each of the clusters identified in cohort B. Next, predicted clusters in cohort A based on the clusters of cohort B were compared to the ‘real’ clusters of cohort A. This was done for each of the three pairwise comparisons (DCS-GoDARTS, DCS-ANDIS, GoDARTS-ANDIS). Agreement between clusters was accessed based on the specificity and sensitivity.

Analyses were performed using R statistics (version 3.6.2). Figures were produced using the R-package *ggplot2* (v3.3.0) and *omicCircos* (v1.22.0).

## RESULTS

### Clustering in three large cohorts based on clinical measures

In this cross-sectional study, 15,940 individuals from three cohorts were included for which baseline characteristics are given in Table 1. The characteristic of the three cohorts were generally comparable with the majority males and an average of around sixty years. Individuals were clustered based on age, BMI, HbA1c, C-peptide and HDL. Based on the gap-statistic across the three cohorts, the most optimal number of clusters was five (Fig. 1, Fig. S1a). The first cluster comprised 13-17% of the individuals included. It was characterized by high HbA1c, but compared to the other clusters, younger with lower BMI, C-peptide and HDL levels. When compared to the original clusters in ANDIS[1], this cluster was most similar to the Severe Insulin-Deficit Diabetes (SIDD) cluster with a sensitivity (SE) of 90.7% (confidence interval (CI), 88.4-92.6%, Fig. 1, Fig. S1b).[1] Between 9 and 22% of individuals clustered to a cluster with high C-peptide levels and age, but relatively lower Hba1c and HDL levels, suggestive of insulin resistance. Indeed, compared to the ANDIS clusters, this cluster resembled most that of the Severe Insulin-Resistant Diabetes cluster (SIRD) with SE of 92.4% (CI, 89.7-94.6%, Fig. 1, Fig. S1b).[1] The third cluster was comprised of participants with high BMI and the youngest age and relatively lower levels of HbA1c and HDL. It was most similar to the originally described Mild Obesity-related Diabetes cluster (MOD) with a SE of 80.6% (CI, 78.4-82.7%) and comprised 18-23% of the individuals included in the study. The fourth and fifth clusters were most similar to the Mild Age Related Diabetes cluster and showed a combined sensitivity of 79.1% (CI, 77.5-80.6%) against the MARD cluster in ANDIS (Fig. 1, Fig. S1b).[1] The fourth cluster, which was also the largest encompassing 29-35% of the individuals, showed no extreme characteristics and was termed Mild Diabetes. The fifth cluster was characterized by higher age and HDL and termed Mild Diabetes with high HDL (MDH) and comprised 16-19% of the individuals, Fig. 1).

**Table 1.** Characteristics of the included individuals of the three cohorts.

| *Variable* | *DCS* | *GoDARTS* | *ANDIS* |
| --- | --- | --- | --- |
| N | 2953 | 5509 | 7478 |
| Males, % | 55.9 | 56.3 | 60.1 |
| Age, years | 60.2[53.1-66.9] | 62.5[54.5-70.0] | 62.0[54.0-69.8] |
| BMI, kg/m<sup>2</sup> | 29.5[26.7-33.2] | 31.0[27.6-35.1] | 30.8[26.9-34.0] |
| HbA1c, mmol/mol | 49.7[44.0-60.7] | 58.0[50.0-79.0] | 62.3[45.0-74.0] |
| HbA1c, % | 6.7[6.2-7.7] | 7.5[6.7-9.4] | 7.9[6.3-8.9] |
| C-peptide, nmol/L | 1.0[0.8-1.4] | 1.9[1.3-2.6] | 1.3[0.9-1.5] |
| HDL, mmol/L | 1.2[0.97-1.37] | 1.1[1.0-1.4] | 1.2[0.9-1.4] |
| LDL, mmol/L | 3.0[2.3-3.7] | 2.7[2.0-3.4] | 3.2[2.5-3.9] |
| Triacylglycerol, mmol/L | 1.7[1.2-2.3] | 2.3[1.6-3.3] | 2.2[1.2-2.4] |
| Glucose lowering, % | 61.5 | 19.0 | 59.6 |

**Figure 1.**
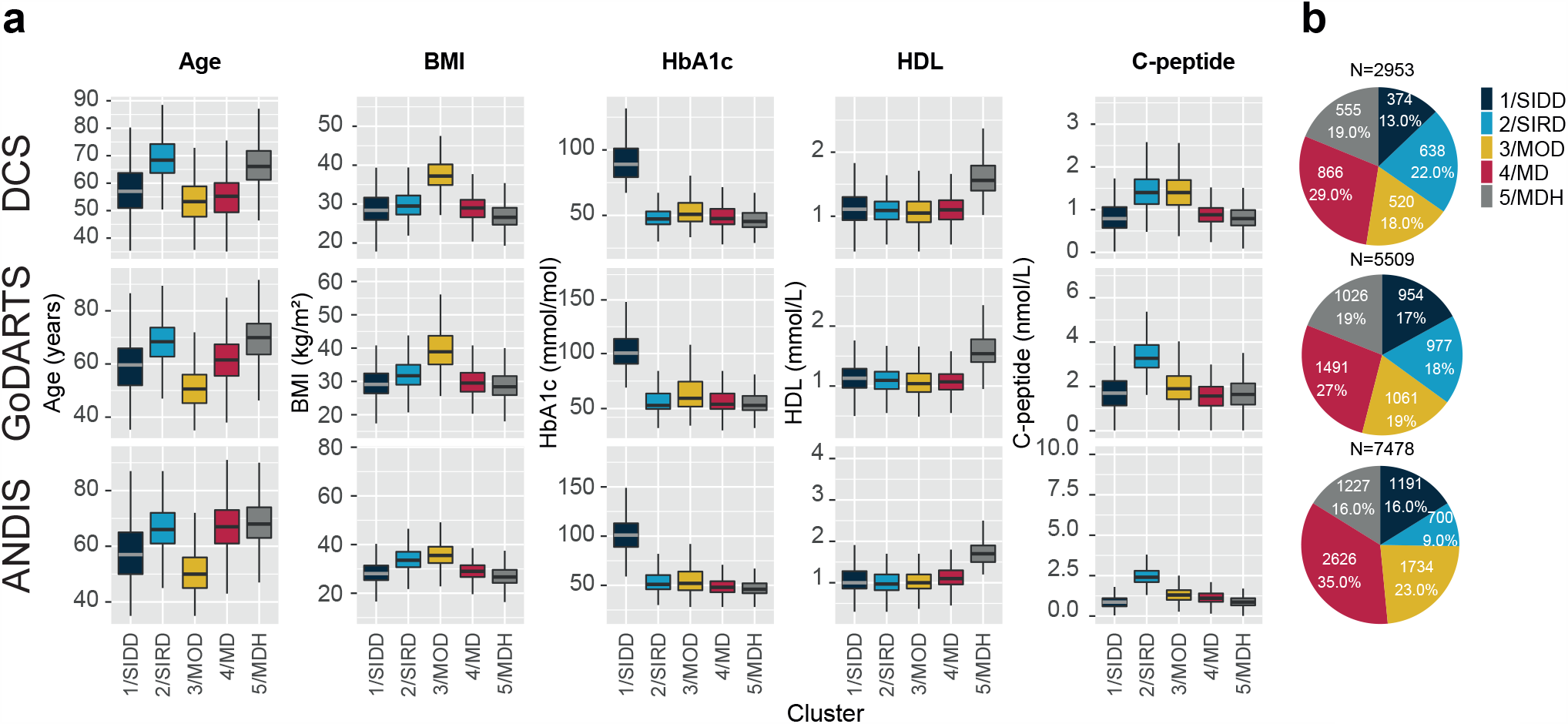
Characteristics of the clusters. **a**. Characteristics of the five clusters across three cohorts DCS, GoDARTS and ANDIS. X-axis, cluster. Y-axis, age, BMI, HbA1c, HDL and C-peptide **b**. Frequency and percentage of individuals in each of the clusters. SIDD, Severe Insulin-Deficit Diabetes; SIRD, Severe Insulin-Resistant Diabetes cluster; MOD, Mild Obesity-related Diabetes; MD, Mild diabetes; MDH, Mild diabetes with high HDL.

### Clusters cross-validate between the three cohorts

To assess the stability across cohorts, clusters were cross-validated between cohorts. Clusters generally cross-validated well between the three cohorts (Fig. S2, Table S1). The SIDD and MDH clusters showed the highest sensitivity of the five clusters identified ranging from 85.6% (CI, 83.5-87.6%) to 97.1% (CI, 94.8-98.5%) and 73.3% (CI, 69.5-77.0%) to 92.9% (CI, 91.3-94.3%, Fig. S2, Table S1). The SIRD and the MD cluster performed generally worst in terms of sensitivity, with sensitivities ranging from 36.1% (CI, 32.3-39.9%) to 92.3% (CI, 90.1-94.2%) in SIRD and 40.8% (CI, 38.9-42.7%) to 78.1% (CI, 75.9-80.2%). Individuals clustered to SIRD were classified as MD and vice versa (Fig. S2, Table S1). The sensitivity of the MOD cluster ranged from 55.0% (CI, 52.6-57.3%) to 93.2% (CI, 91.5-94.7%).

## DISCUSSION

Based on five clinical variables, people with type 2 diabetes from three large European cohorts were assigned to five separate clusters. Clusters were successfully cross-validated against the clustering reported by Ahlqvist et al.[1] but also between cohorts included.

Even though we used slightly different parameters for clustering – i.e. C-peptide and HDL instead of HOMA measures [1], people were clustered largely to the same clusters in a direct comparison with previously published clusters in ANDIS. The insulin deficient cluster (SIDD) was defined by a high HbA1c, the insulin resistant by a high C-peptide (SIRD), and the obese cluster by a high BMI (MOD). The previously identified MARD cluster [1], could be further divided into two clusters of people with a low (MD-cluster) and a high HDL (MDH-cluster). Including HDL resulted two clusters with mild characteristics, where one had high HDL and one cluster generally a low HLD. A subset of the SIRD cluster was classified as MD, which is most likely due to the use of C-peptide and HDL instead of HOMA measures.

In addition to a comparison to the original ANDIS clusters, in the current study we also cross-validated the clusters across cohorts. Clusters cross-validated generally well and the best sensitivity was observed in the SIDD and MDH clusters. For SIRD and MD a lower sensitivity was observed. Individuals that were classified in one cohort to SIRD but also MOD, were classified as MD in a second cohort and vice versa. The characteristics of particularly SIRD and MD are very similar with the sole difference higher levels of C-peptide in the SIRD cluster. This could explain the difference in classification in the two cohorts.

## CONCLUSION

In the current study, clusters were identified in three cohorts, based on five different clinical characteristics. We show that clusters based on random or fasted C-peptide instead of HOMA measures result in clusters that resemble those based on HOMA measures. By adding HDL we identified one additional cluster with mild characteristics. Cross-validation between cohorts showed that there was generally a good resemblance between cohorts. Together, our results show that the clustering is generally stable across cohorts, also when the clustering includes C-peptide instead of HOMA measures.

## Data Availability

Clinical data will not be deposited for this study.

## ACKNOWLEDGEMENTS

This project has received funding from the Innovative Medicines Initiative 2 Joint Undertaking under grant agreement No 115881 (RHAPSODY). This Joint Undertaking receives support from the European Union’s Horizon 2020 research and innovation programme and EFPIA. This work is supported by the Swiss State Secretariat for Education, Research and Innovation (SERI) under contract number 16.0097-2. The opinions expressed and arguments employed herein do not necessarily reflect the official views of these funding bodies. We acknowledge the support of the Health Informatics Centre, University of Dundee for managing and supplying the anonymised data. ERP was supported by a Wellcome Trust investigator award (102820/Z/13/Z). GAR was supported by a Wellcome Trust Senior Investigator (WT098424AIA) and Investigator Award (212625/Z/18/Z), MRC Programme grants (MR/R022259/1, MR/J0003042/1, MR/L020149/1) and by Diabetes UK (BDA/11/0004210, BDA/15/0005275, BDA 16/0005485) project grants.

## AUTHOR CONTRIBUTIONS

RCS, LAD, JWJB, LMTH, ERP designed the study and drafted the manuscript. RCS, LAD, HF, GAB, MA performed the analyses. ID, DK, MI set up a federated node system for data-analysis. RCS, DMA, LAD, HF, EA, AA, MJG, MK, FM, TS, AW, CLQ, MI were involved in the data pre-processing and quality control. GNG, AF, MKH, DMA, IP, TJP, BT, VL, LG, PWF, GAR, MJG, CK, KS, CLQ, AA, PR, AW, TS contributed to the data acquisition and project logistics. All authors contributed to the data interpretation. All authors critically revised the manuscript and approved the final version. RCS, LAD, JWJB, LMTH, ERP are the guarantors of the work.

## SUPPLEMENTARY FIGURES

**Figure S1 GAP statistic and comparison to clusters based on HOMA measures a.** Number of clusters versus the GAP statistic in DCS, GoDARTS and ANDIS. The most optimal number of clusters was at five clusters indicated by the dotted line. **b**. Comparison of clustering in ANDIS in the current study versus those identified in Ahlqvist et al.[1]. Numbers on the diagonal represent the absolute overlap (upper number) and the percentage (lower number). In the right panel, the sensitivity and specificity are given with 95% confidence intervals. SIDD, Severe Insulin-Deficit Diabetes; SIRD, Severe Insulin-Resistant Diabetes cluster; MOD, Mild Obesity-related Diabetes; MD, Mild diabetes; MDH, Mild diabetes with high HDL.

**Figure S2 All pairwise comparisons of identified clusters versus predicted clusters.** Top number in each cell represents the absolute overlap, the bottom number the percentage of individuals. Individuals were assigned to a cluster in cohort A based on the cluster centres of cohort B and compared to the clusters identified in the cohort A. This resulted in the following combinations: **a** clusters identified in DCS based on ANDIS centres **b** clusters identified in ANDIS based on DCS centres **c** clusters identified in ANDIS based on GoDARTS centres **d** clusters identified in GoDARTS based on ANDIS centres **e** clusters identified in GoDARTS based on DCS centres **f** clusters identified in DCS based on GoDARTS centres. SIDD, Severe Insulin-Deficit Diabetes; SIRD, Severe Insulin-Resistant Diabetes cluster; MOD, Mild Obesity-related Diabetes; MD, Mild diabetes; MDH, Mild diabetes with high HDL.

## SUPPLEMENTARY TABLES

**Table S1.** Sensitivity and specificity of the cross-validation of clusters in other studies.

| Group | Sensitivity | Specificity | Cohort | Centers |
| --- | --- | --- | --- | --- |
| 1/SIDD | 96.22[94.98-97.23] | 98.62[98.3-98.89] | GoDARTS | ANDIS |
| 1/SIDD | 97.06[94.8-98.52] | 97.32[96.63-97.91] | ANDIS | DCS |
| 1/SIDD | 85.64[83.52-87.59] | 99.63[99.45-99.77] | DCS | ANDIS |
| 1/SIDD | 88.78[86.61-90.72] | 98.59[98.21-98.92] | ANDIS | GoDARTS |
| 1/SIDD | 77.57[74.79-80.18] | 99.19[98.88-99.43] | DCS | GoDARTS |
| 1/SIDD | 97.06[94.8-98.52] | 96.36[95.56-97.04] | GoDARTS | DCS |
| 2/SIRD | 92.29[90.05-94.15] | 92[91.33-92.64] | GoDARTS | ANDIS |
| 2/SIRD | 36.05[32.32-39.91] | 94.43[93.41-95.33] | ANDIS | DCS |
| 2/SIRD | 68.57[64.99-72] | 82.75[81.83-83.65] | DCS | ANDIS |
| 2/SIRD | 56.19[53.02-59.33] | 99.29[99-99.52] | ANDIS | GoDARTS |
| 2/SIRD | 83.42[80.94-85.7] | 89.67[88.75-90.54] | DCS | GoDARTS |
| 2/SIRD | 64.26[60.41-67.99] | 94.25[93.23-95.17] | GoDARTS | DCS |
| 3/MOD | 76.01[73.93-78] | 98.45[98.1-98.75] | GoDARTS | ANDIS |
| 3/MOD | 73.85[69.84-77.57] | 85.16[83.69-86.55] | ANDIS | DCS |
| 3/MOD | 54.96[52.58-57.32] | 95.73[95.18-96.24] | DCS | ANDIS |
| 3/MOD | 93.21[91.53-94.65] | 92.72[91.91-93.46] | ANDIS | GoDARTS |
| 3/MOD | 62.87[59.88-65.78] | 96.27[95.67-96.81] | DCS | GoDARTS |
| 3/MOD | 74.23[70.24-77.94] | 91.62[90.44-92.69] | GoDARTS | DCS |
| 4/MD | 66.15[64.3-67.96] | 91.53[90.71-92.3] | GoDARTS | ANDIS |
| 4/MD | 51.04[47.65-54.42] | 74.46[72.53-76.32] | ANDIS | DCS |
| 4/MD | 40.82[38.93-42.73] | 81.04[79.91-82.13] | DCS | ANDIS |
| 4/MD | 78.14[75.95-80.21] | 83.13[81.93-84.27] | ANDIS | GoDARTS |
| 4/MD | 67.81[65.37-70.17] | 86.96[85.88-87.99] | DCS | GoDARTS |
| 4/MD | 66.74[63.5-69.88] | 87.83[86.35-89.2] | GoDARTS | DCS |
| 5/MDH | 92.75[91.15-94.13] | 94.18[93.57-94.74] | GoDARTS | ANDIS |
| 5/MDH | 73.33[69.45-76.97] | 98.54[97.98-98.98] | ANDIS | DCS |
| 5/MDH | 92.91[91.33-94.28] | 92.71[92.03-93.34] | DCS | ANDIS |
| 5/MDH | 76.02[73.29-78.61] | 98.19[97.76-98.56] | ANDIS | GoDARTS |
| 5/MDH | 89.47[87.43-91.28] | 96.36[95.77-96.89] | DCS | GoDARTS |
| 5/MDH | 85.41[82.19-88.24] | 97.62[96.93-98.19] | GoDARTS | DCS |

*SIDD, Severe Insulin-Deficit Diabetes; SIRD, Severe Insulin-Resistant Diabetes cluster; MOD, Mild Obesity-*
*related Diabetes; MD, Mild diabetes; MDH, Mild diabetes with high HDL.*

