## Supplementary figures and images for "Replication and cross-validation of T2D subtypes based on clinical variables: an IMI-RHAPSODY study"

### Fig. S1

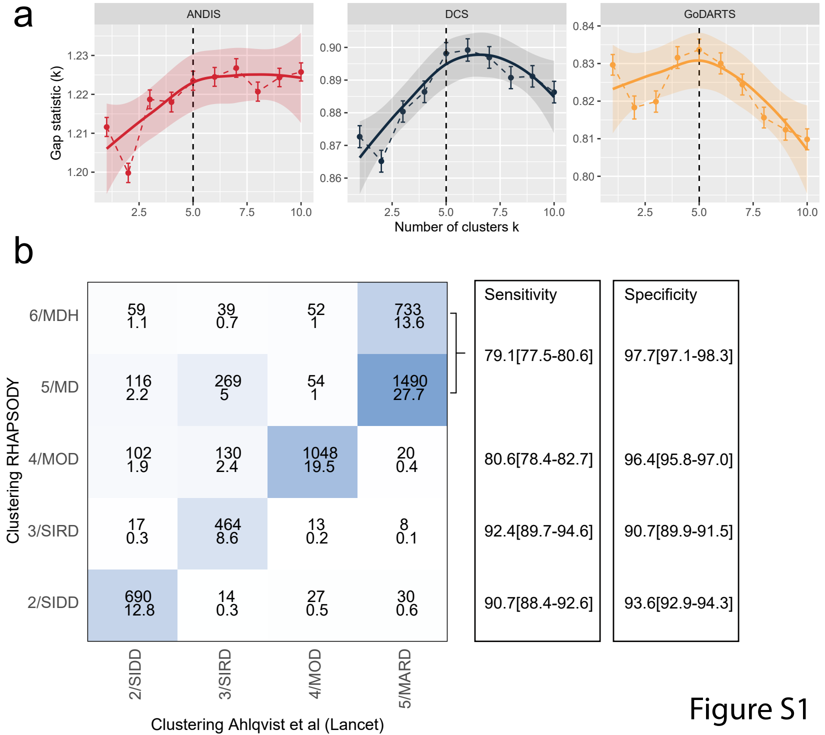

### Fig. S2

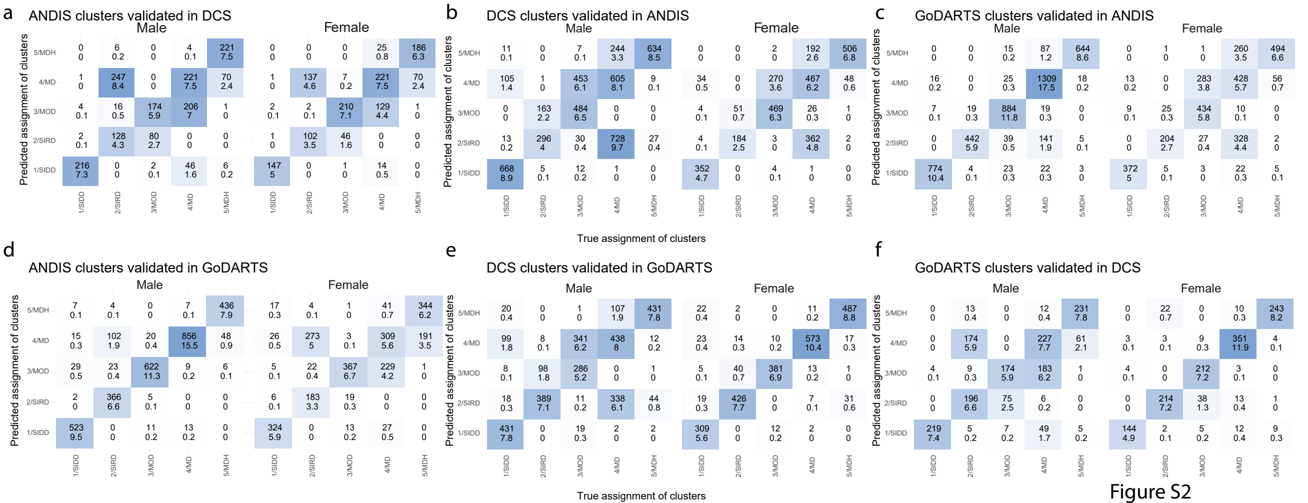
